# Toward Evaluation of Disseminated Effects of Medications for Opioid Use Disorder within Provider-Based Clusters Using Routinely-Collected Health Data

**DOI:** 10.1101/2022.04.15.22273847

**Authors:** Ashley L. Buchanan, Tianyu Sun, Jing Wu, Hilary Aroke, Jeffrey Bratberg, Josiah Rich, Stephen Kogut, Joseph Hogan

## Abstract

Routinely-collected health data can be employed to emulate a target trial when randomized trial data are not available. Patients within provider-based clusters likely exert and share influence on each other’s treatment preferences and subsequent health outcomes and this is known as dissemination or spillover. Extending a framework to replicate an idealized two-stage randomized trial using routinely-collected health data, an evaluation of disseminated effects within provider-based clusters is possible. In this paper, we propose a novel application of causal inference methods for dissemination to retrospective cohort studies in administrative claims data and evaluate the impact of the normality of the random effects distribution for the cluster-level propensity score on estimation of the causal parameters. An extensive simulation study was conducted to study the robustness of the methods under different distributions of the random effects. We applied these methods to evaluate baseline prescription for medications for opioid use disorder among a cohort of patients diagnosed opioid use disorder and adjust for baseline confounders using information obtained from an administrative claims database. We discuss future research directions in this setting to better address unmeasured confounding in the presence of disseminated effects.

## 1 Introduction

Routinely-collected health data, such as electronic health records and administrative claims, can be employed to emulate a target trial when randomized trial data are not available due to ethical or financial constraints. Patients within provider-based clusters could exert influence on each other’s treatment preferences and subsequent health outcomes possibly due to shared prescription medications, known as medication diversion [1, 2]. In addition, there is shared influence on patients’ treatments due to shared providers and geographical proximity, which could impact social norms around prescription utilization and medical treatment [3–6]. In the literature, this is known as spillover, interference, or dissemination [7–9]. There is evidence of clustering of prescribing patterns by practice types, including variation in prescription refills and duration by patient type [10] and provider practice[11, 12], as well as recent studies documenting medication diversion [13, 14].

We define a *provider cluster* to be all patients with the diagnosis of interest seen at least once by particular provider during a specified time period. We will refer to the patients who were prescribed medications of interest as the *treated* patients and those who are not, but shared a provider cluster with the treated patients as *untreated* patients. *Coverage* of a prescription treatment is defined as the proportion of patients who are prescribed the medication(s) of interest by a particular provider during a specified time period. The *direct* effect is the difference in average potential outcomes under MOUD prescription versus no MOUD prescription with a fixed coverage level of MOUD. The *disseminated* (i.e., indirect or spillover) effect is the difference in average potential outcomes of an untreated patient (i.e., patients who shared a provider that prescribed the medication, but were not prescribed the medications themselves) under two different coverage levels (Figure 1). Broadly, dissemination (interference or spillover) is when one patient’s exposure affects another patient’s outcome [8]. There are many different mechanisms by which this can occur [15], but in this work, we remain agnostic to the specific pathway. Understanding these effects yields new information to develop and improve provider- and patient-level interventions that could have substantial resonance beyond the patient who was prescribed the medication. Ignoring disseminated effects can generate misleading results, often underestimating the full impact of interventions [16]. To note, the estimands considered in this context are different from similar terminology used for evaluation of mediation (e.g., natural direct and indirect effects) [17].

**Figure 1:**
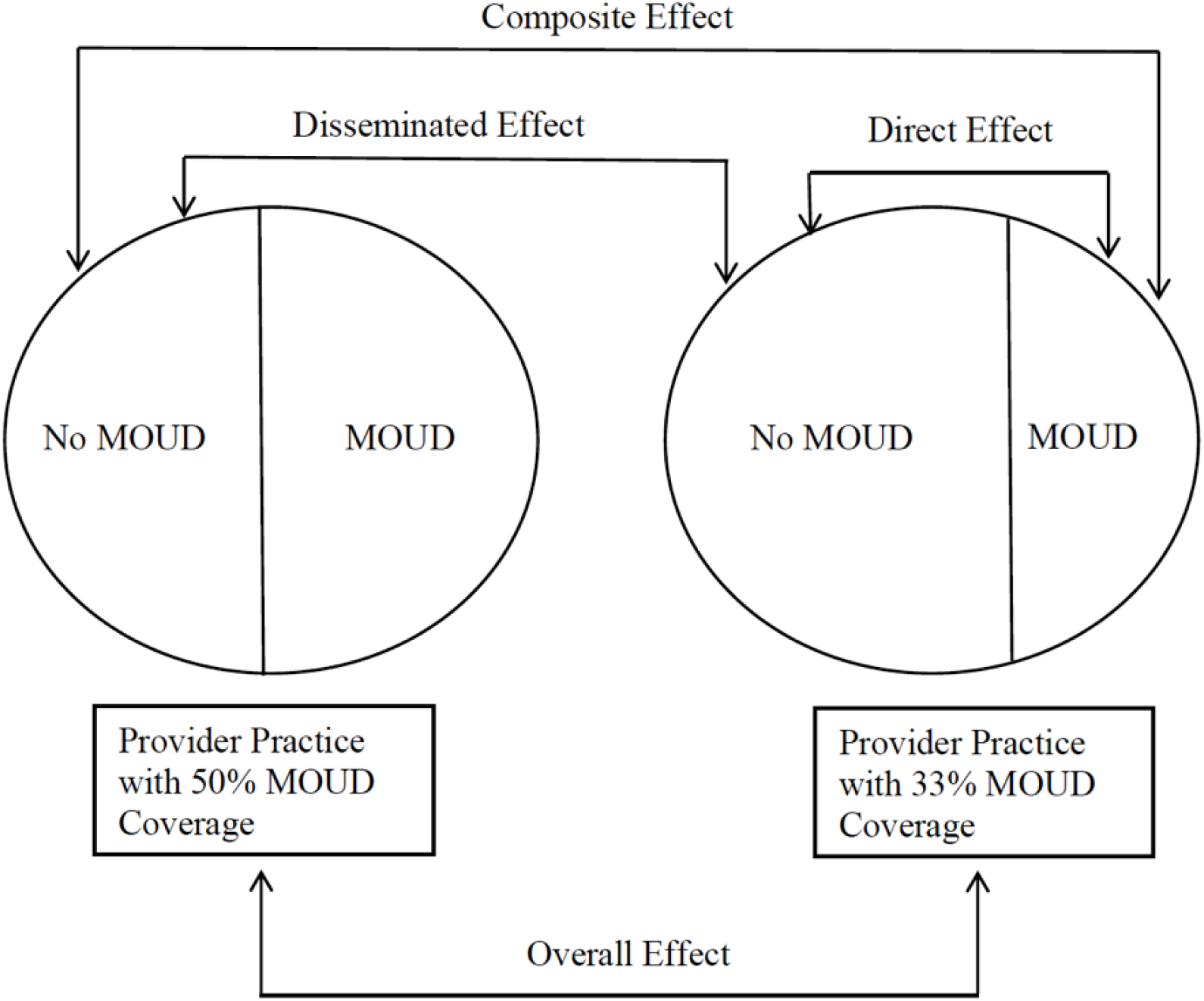
Schematic diagram of the subsets of data used for each estimator (direct, disseminated, composite, and overall) based on a format provided in Halloran and Struchiner [79]

Most current methodology in the presence of disseminated effects may require extensions for studies in large administrative claims databases. Existing methodology was developed for randomized or observational cohort study designs; whereas, studies in administrative claims databases are often retrospective designs primarily concerned with effectiveness and cost-effectiveness and face unique methodological challenges [18]. Available estimators for two-stage randomized trials or observational studies with an outcome at a single time point often rely on the assumption of no dissemination between study clusters [7, 19]. Recent work provides a conceptual framework for evaluating disseminated effects using routinely-collected health data [20].

An important assumption in application of this approach is the normality of the random effects. Generalized linear mixed effects models with random effects included in the linear predictor can be employed for inference in clustered data. In a linear mixed effects model, deviations from the normality assumption of the random effects will not typically impact the inference about the fixed effects used for prediction [21, 22]. For the estimators of disseminated effects, we assume the random effects are asymptotically normally distributed. Then, this distribution is employed to integrate over the random effects for each cluster to obtain the joint probability of treatment in a provider cluster. In this case, the distributional assumption about the random effects could be more critical to ensure consistent inference, even if the fixed effects are correctly specified, but this has not been fully addressed yet in the literature.

In this work, we propose a novel application of causal inference methods to evaluate disseminated effects in retrospective studies using administrative claims data and discuss the assumptions needed to identify causal effects. The methodological contribution of this paper is an empirical investigation into the robustness of the assumptions about the cluster-level propensity score model when evaluating disseminated effects. With limited theoretical results for the impact of misspecification on these methods, an empirical investigation can provide new insights and improve recommendations for future applications. This paper contributes to our understanding of how to employ inverse probability weighted estimators to quantify dissemination in administrative claims data, which requires careful considerations about assumptions and the construction of the data set, closely following a trial emulation approach [23]. We describe an extensive simulation study to study the robustness of the methods under different distributions of the random effects in the treatment propensity score model. We apply these methods to evaluate the effectiveness of baseline prescriptions for medications for opioid use disorder to prevent overdose among a cohort of patients with diagnosed opioid use disorder and adjust for baseline confounders using information from an administrative claims database. We consider the direct effect of being prescribed medications with a fixed coverage in the provider cluster, as well as the disseminated effect of sharing a provider that prescribed the medications, but not being prescribed medications themselves under two different coverage levels. Lastly, we discuss future research directions for retrospective studies in the presence of dissemination.

### 1.1 Motivating Example

Medications for opioid use disorder (MOUD) show great promise in curtailing the opioid crisis in the United States [24]. Treatment of opioid use disorder (OUD) is a huge burden on the medical system and understanding behavioral and social norms to improve the uptake of MOUD could alleviate some of this burden. MOUD is a potentially life-saving intervention for patients with OUD and typically includes a prescription for either buprenorphine/naloxone, methadone, or naltrexone [25–27], along with behavioral therapy. However, treatment access to MOUD for those with OUD remains limited. Nationally in 2019, less than 20% of people with OUD received medication for OUD [28]. Increasing efforts have been made to encourage the uptake of buprenorphine/naloxone as treatment for patients facing OUD, including telehealth initiation of treatment, expansion of Drug Enforcement Agency (DEA) waivers, and dispensing at community pharmacies [29–33].

Earlier studies have considered rates of MOUD prescribing from both a patient and provider perspective [34, 35], as well as the effects of MOUD on subsequent health outcomes [36, 37]; however, few studies have evaluated the possible disseminated effects of MOUD on overdose prevention among patients diagnosed with OUD. There is documented evidence of medication diversion in this patient population [13, 14]. Sharing buprenorphine for detoxification or reducing withdrawal symptoms was common among patients prescribed buprenorphine for OUD with about 50% reporting sharing their prescription with others [38] and patients who reported using diverted medications had an associated reduction in use of illicit substances [39]. Although patient sharing of MOUD is not itself a solution to the undertreatment of patients with OUD, understanding the effects of MOUD within provider clusters can provide insights into MOUD coverage levels required to improve outcomes among people diagnoised with OUD. To evaluate disseminated effects in provider-based clusters, we defined fixed clusters according to providers (e.g., most frequently seen provider) and assumed that dissemination occurs within, but not between provider-based clusters, which is known in the literature as the *partial interference* assumption [40].

Among patients diagnosed with OUD, we evaluated the direct effect of MOUD for a fixed coverage in the provider cluster and also the disseminated effect to untreated patients (those sharing a provider-based cluster with those who were prescribed MOUD, but not prescribed MOUD themselves) using an administrative health claims database. Optum’s de-identified Clinformatics® Data Mart Database is a commercial and Medicare Advantage claims database [41]. The statistically de-identified data includes medical and pharmacy claims, as well as laboratory results, from 2010 through 2015 with over 35 million enrollees from birth to 65+ years. The data are blinded to protect patient privacy and included the following patient-linked longitudinal data: demographics on the member, physician and facility claims data, pharmacy claims data, laboratory test results, inpatient data, medication and procedural cost information.

## 2 Assumptions and Notation

Patients entered this study at the time of their first observed OUD diagnosis in the routinely-collected database (Figure 2). A fixed time period directly preceding the OUD diagnosis was used to define the *baseline period*, while a fixed period immediately after the OUD diagnosis, possibly 90 days, was used to define variables related to MOUD initiation and initial exposure status, referred to as the *index period*. After the index period, patients were followed for a fixed duration to ascertain incident outcomes of interest with available information and patient follow-up ends when the outcome occurs, administrative censoring, or the patient leaves the database due to death or disenrollment. Following a new user design, we excluded people who were prescribed MOUD during baseline and we also excluded people who had opioid overdose during the baseline or index periods [42]. This new user design allows for alignment with the conceptualization of this study as a randomized trial, except in this setting the treatment was not randomized. This design also protects against biases of a prevalent user design, specifically missed events that occur early in treatment and baseline covariates themselves that may be associated with prior treatment.

**Figure 2.**
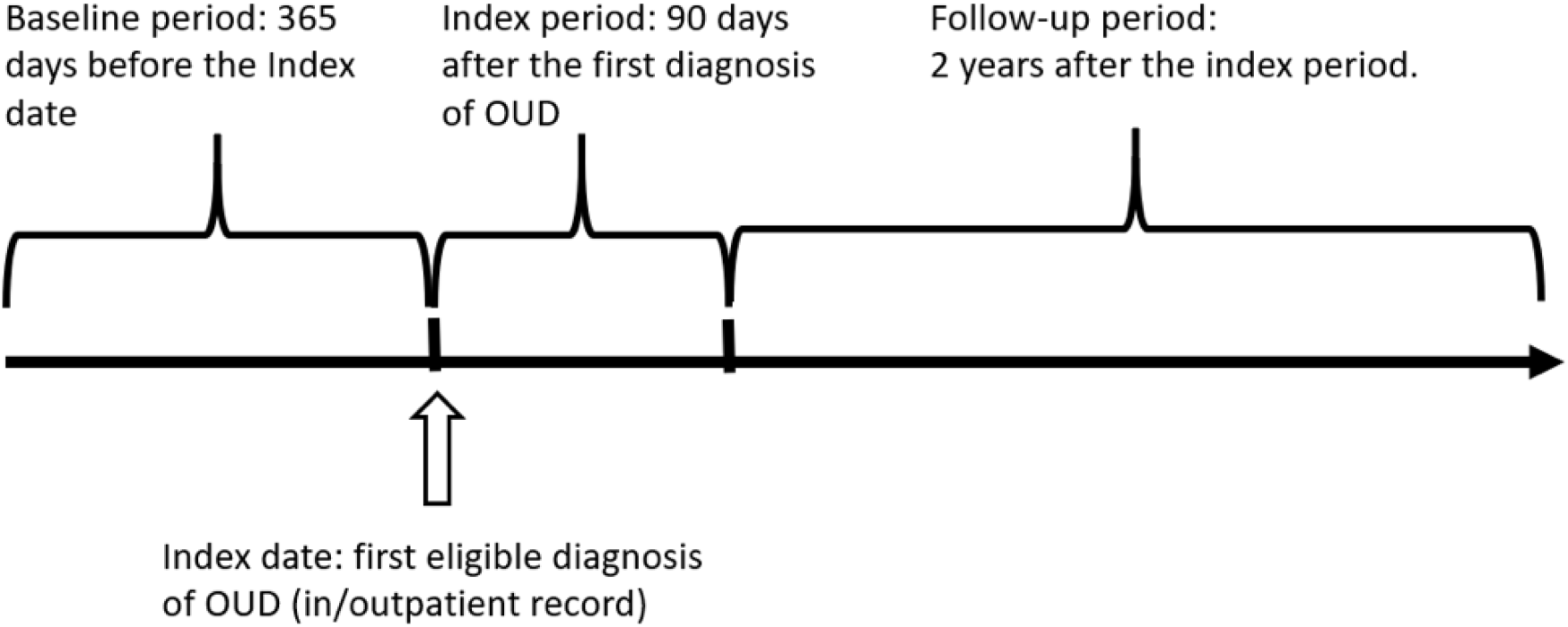
: Schematic diagram of retrospective cohort study of medications for opioid use disorder (MOUD) on the two-year risk of opioid overdose among patients diagnosed with opioid use disorder (OUD) in Optum’s de-identified Clinformatics® Data Mart Database, 2010-2017, United States^*a*^

There are *K* provider-based clusters (*k* = 1, …, *K*) with *i* = 1, …, *n*_*k*_ patients in each provider cluster with the total number of patients denoted by 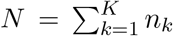. Define 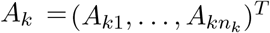 as the vector of baseline *treatment indicators* for patients in cluster *k* and let 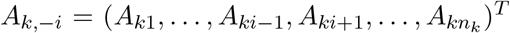. In this setting, *A*_*ki*_ corresponds to an indicator of the receipt of a prescription for MOUD and no information was available on adherence to MOUD. the Let *y*_*ki*_(*a*_*ki*_, *a*_*k,−i*_) denote the potential outcome of patient *i* if they received treatment *a*_*ki*_ and their provider cluster had assignment *a*_*k*_ with *a*_*k,−i*_ as the treatment indicators for other patients in the cluster. We can also write this potential outcome as *y*_*ki*_(*a*_*k*_). By consistency, *Y*_*ki*_ = *y*_*ki*_(*A*_*k*_) is the observed outcome. To ensure a temporal sequence between treatment and outcome, the assessment of outcome occurred after receipt of the prescription, specifically after the period used to define the prescription exposure (i.e., the index period). In this approach, we considered the first incident outcome within a defined length of follow-up after the index period. We assumed that the distribution of counterfactual treatment under a Bernoulli allocation strategy defined the counterfactual estimands of interest. This can be conceptualized as standardizing to a randomized trial in which treatment was assigned according to this strategy. Let *α* correspond to a counterfactual scenario where patients independently receive treatment with probability *α*. In the motivating example, the counterfactual allocation strategies *α* correspond to different coverage levels of MOUD assigned to each cluster, then patients are randomized according to a given allocation strategy within the cluster. Let *X*_*ki*_ denote pre-treatment individual level covariates for patient *i* in cluster *k* and *X*_*k*_ denote a vector of pre-treatment cluster level covariates among all patients in cluster *k*.

We consider the disseminated effects of MOUD prescriptions to untreated patients received through exposure to the non-randomized MOUD prescription provided only to the treated patients in each cluster. For example, patients may share MOUD prescriptions with each other [1, 2, 29] or may influence each other to seek OUD treatment either from a medical provider, methadone clinic, or other settings [3–6]. We conducted a retrospective observational study and were interested in the intention-to-treat effects, assuming noninformative dropout from the database (e.g., loss of insurance eligibility). The sufficient conditions for estimating valid effects have been previously described, including: (1) representativeness of the unexposed for the treatment response had they been exposed and vice versa conditional on baseline covariates (i.e., conditional exchangeability); (2) homogeneity of treatment effects despite any variations that may occur in practice, and no multiple versions of treatment [43–46]; (3) no measurement error in any variable needed for valid analysis; and (4) no dissemination between provider-based clusters [47, 48]; (5) cluster-level positivity assumption for the propensity score[47].

Patients who are prescribed MOUD are likely different from patients who are not prescribed as part of their OUD care, specifically patients prescribed MOUD may be later in their disease or treatment trajectory with an associated increased risk of a history heroin/fentanyl or injection drug use [49–52] Furthermore, providers who prescribe MOUD at different rates likely differ in their approach to pain management and identification and treatment of OUD [12, 53]. Dissemination within a provider cluster is of primary interest in this setting and conditioning on a set of pre-treatment covariates is assumed to be sufficient to control for confounding. That is, the potential outcomes of those who were exposed and the potential outcomes of those who were not exposed are assumed to be the same on average conditional on observed baseline covariates (i.e., baseline conditional exchangeability). We also make the stratified interference assumption, which assumes that an patient’s potential outcome is dependent only on their own treatment and the proportion of those treated among all patients seen by their provider. In addition, we assume partial interference defined by provider practices; that is, we assume potential outcomes depend only on treatments of patients within a provider cluster and not on treatments (or outcomes) of others [40]. We assume that if a patient was prescribed MOUD that they filled that prescription. This approach also requires a positivity assumption for the cluster-level propensity score; that is, a positive probability of the cluster-level treatment conditional on the covariates [47].

## 3 Causal Framework and Parameters

Conceptualizing a study as a randomized trial can improve information gleaned from observational studies (Table 1) [23]. Because it is unethical to withhold treatment for patients with opioid use disorder and MOUD has already demonstrated effectiveness in randomized trials, observational data analysis can be used to evaluate possible disseminated effects of MOUD. Furthermore, a two-stage randomized design, in which provider clusters are first randomized to a MOUD allocation strategy (e.g., 50% of patients for a given provider are prescribed MOUD), then patients within the provider practice are randomized to MOUD (or not) according to that strategy, would be difficult to implement. Providers have treatment preferences and may not be amenable to an assigned MOUD coverage level in their practice. In this target trial design, we estimate intention-to-treat effects using a comparison of two-year opioid overdose risks among patients assigned to MOUD at baseline, compared to those assigned to no MOUD at baseline. When we emulate this design using observational data, we use the receipt of MOUD prescription as a proxy for assignment and, due to the lack of randomization, appropriate adjustment for confounding is required.

**Table 1:**
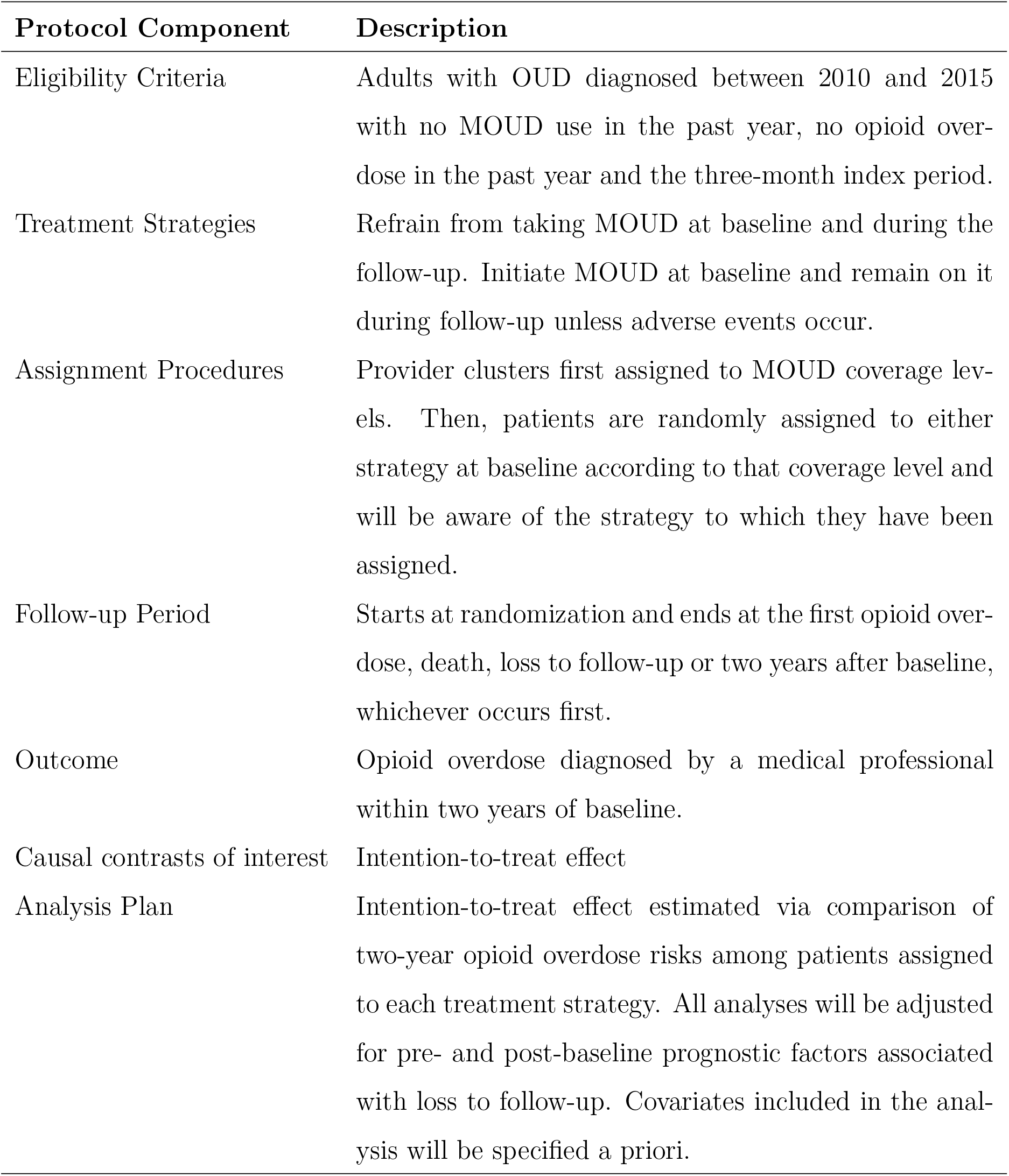
Protocol of a target trial to estimate the effect of medications for opioid use disorder (MOUD) on the two-year risk of opioid overdose among those with opioid use disorder

| Protocol Component | Description |
| --- | --- |
| Eligibility Criteria | Adults with OUD diagnosed between 2010 and 2015 with no MOUD use in the past year, no opioid overdose in the past year and the three-month index period. |
| Treatment Strategies | Refrain from taking MOUD at baseline and during the follow-up. Initiate MOUD at baseline and remain on it during follow-up unless adverse events occur. |
| Assignment Procedures | Provider clusters first assigned to MOUD coverage levels. Then, patients are randomly assigned to either strategy at baseline according to that coverage level and will be aware of the strategy to which they have been assigned. |
| Follow-up Period | Starts at randomization and ends at the first opioid overdose, death, loss to follow-up or two years after baseline, whichever occurs first. |
| Outcome | Opioid overdose diagnosed by a medical professional within two years of baseline. |
| Causal contrasts of interest | Intention-to-treat effect |
| Analysis Plan | Intention-to-treat effect estimated via comparison of two-year opioid overdose risks among patients assigned to each treatment strategy. All analyses will be adjusted for pre- and post-baseline prognostic factors associated with loss to follow-up. Covariates included in the analysis will be specified a priori. |

The *direct* effect is the difference in average potential outcomes under MOUD versus no MOUD with a fixed coverage level of MOUD prescribing in the prescriber practices [7]. The *disseminated* effect is the difference in average potential outcomes when a patient is untreated under two different coverage levels. This is a measure of the indirect or “spillover” effect of MOUD prescriptions to other patients within the provider-based cluster. Although the methods are agnostic to the exact mechanism, possible “spillover” mechanisms in this setting could include medication diversion [1, 2, 29], and geographical proximity, which could impact social norms around prescription utilization [3–6] and medical treatment [10–12]. The composite effect is the difference in average potential outcomes for treated patients under high MOUD coverage versus untreated patients under low MOUD coverage, and represents the maximal effect of MOUD on patient health outcomes. The overall effect is the difference in average potential outcomes under high coverage versus low coverage, and will change, in general, as the proportion of treated patients (i.e., those prescribed MOUD) in the provider clusters varies [16]. The population average counterfactuals are defined as follows based on the individual average potential outcome; that is, as a function of both the provider cluster treatment strategy assignment and the individual patient’s treatment assignment allocated according to the strategy assigned to the provider. Let 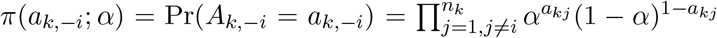 and 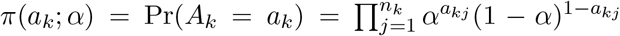. Define 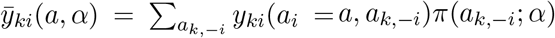 [47, 54]. In words, this is a weighted average of patient *i*’s potential outcomes under all possible treatment vectors of the other *n*_*k*_ *−* 1 patients in cluster *k* weighted by the probability of each possible treatment vector. In this approach, we are standardizing to a setting where the individual treatment allocations are independent and randomly assigned with equal probability, which may be plausible given that the provider prescribes the medication to each patient individually. However, alternative allocation strategies conditional on patient or provider-level covariates could be considered, particularly if information is available on the influence of clinic policies and standards on prescriber practices [55]. Averaging over all patients in each cluster, then over all clusters, we define the population average potential outcome as 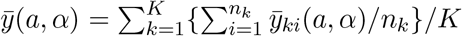. We can also define average potential outcomes only as a function of *α*. Define the marginal average potential outcome for patient *i* under allocation strategy *α* by 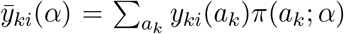. In this case, we take the weighted average of patient *i*’s potential outcomes across all treatment vectors in the cluster. Averaging over patients within each cluster, then over all clusters, define the population average potential outcome as 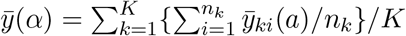.

The direct effect is defined as 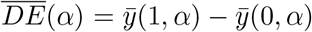, the disseminated (i.e., indirect or spillover) effect is defined as 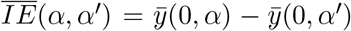, the composite (i.e., total) effect is defined as 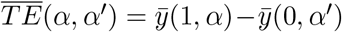), and the overall effect is 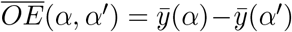, where *α*^*′*^ *< α*. The estimands represent counterfactual scenarios where, for example, treatments are allocated to patients according to an independent Bernoulli distribution with equal probability *α* for each patient within a provider cluster.

## 4 Estimation

We consider both unstabilized Horvitz-Thompson-type and stabilized Hájek-type estimators for the direct, disseminated, composite, and overall effects. If the treatment propensity scores are known, then the effect estimators below are asymptotically unbiased [56]. Alternatively, if the individual propensity scores are unknown, as typically the case with observational studies, we could estimate these treatment propensity scores using a mixed effect logistic regression model logit*{Pr*(*A*_*ki*_ = 1|*X*_*ki*_, *b*_*k*_)*}* = *γ*_0_ + *γ*_**1**_*X*_*ki*_ + *b*_*k*_, where *b*_*k*_ is a random effect to account for clustering within the provider cluster *k* [56]. The joint probability of the treatment in a provider cluster is modeled as

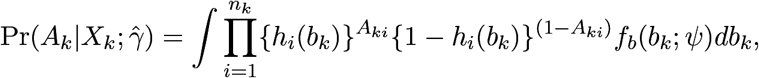

where *h*_*i*_(*b*_*k*_) = Pr(*A*_*ki*_ = 1|*X*_*ki*_, *b*_*k*_) is the probability of treatment conditional on covariates *X*_*ki*_ for patient *i* in cluster *k* and a random effect *b*_*k*_ for cluster *k, f*_*b*_(*b*_*k*_; *ψ*) denotes the density function of *b*_*k*_ which is assumed to be *b*_*k*_ *∼ N* (0, *ψ*). Logistic regression tends to perform well even when the treatment assignment mechanism is unknown [57] and for clustered data, a mixed effects model is a natural way to capture cluster-specific effects for estimation purposes [21]. This second property is appealing in the context of studying dissemination. Using inverse probability weights [47], estimators of the average potential outcomes are

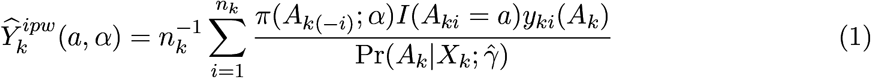

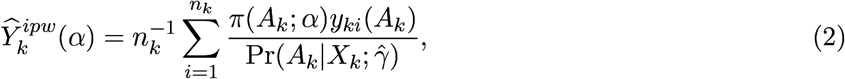

where 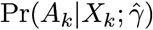 is a model-based estimator of the cluster-level propensity score, e.g., using mixed effects logistic regression. This estimator inverse weights each patient’s outcome by the estimated probability of the provider’s treatment allocation given the covariates of the provider’s patients.

To obtain population averages, define 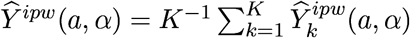 and 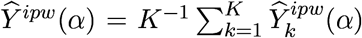. We consider the following risk difference estimators of the direct, disseminated (indirect), composite (total), and overall effects: 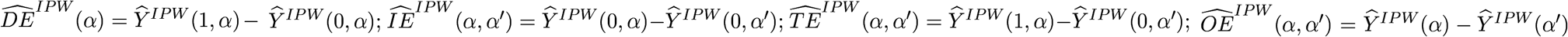. These IPW estimators are consistent and asymptotically normal provided that the treatment propensity score model is correctly specified [54].

We also consider stabilized Hájek-type estimators of the direct, disseminated, composite, and overall effects, which replaces the *n*_*k*_ terms in (1) and (2) with an unbiased estimator. The use of a stabilized estimator can reduce the variance relative to the Horvitz-Thompson type estimator [56]. Let 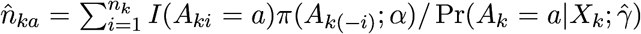. Note that 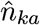 is an unbiased estimators of *n* even in the presence of interference. As proposed in Liu (2016) [56], the following is an alternative estimator for the population average outcome for the treatment *a* and allocation strategy *α*

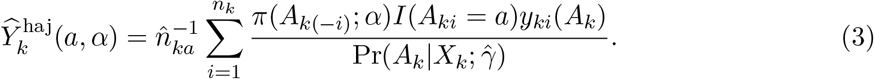

Similarly, let 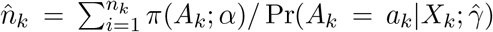. As proposed in Liu (2016) [56], the following is an alternative estimator of the marginal population average outcome for allocation strategy *α*

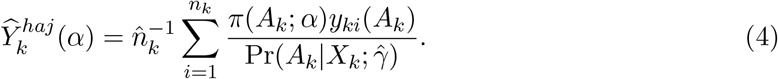

To obtain population averages, define 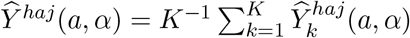 and 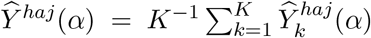. We consider the following risk difference estimators of the direct, disseminated (indirect), composite (total), and overall effects: 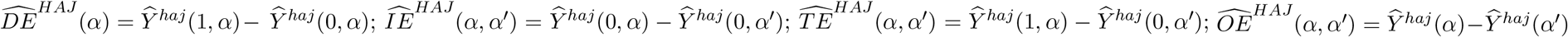. These estimators are consistent and asymptotically normal provided that the treatment propensity score model is correctly specified [54, 56]. We used a robust sandwich-type estimator of the variance to compute Wald-type confidence intervals for these effects [54, 56]. All of the estimators of the direct, disseminated, composite and overall can be analogously defined on the ratio scale.

For the estimators described above, we assumed the random effects were asymptotically normally distributed and validity of the inference requires that the cluster-level propensity score model is correctly specified. The cluster-level propensity score estimator integrates over the distribution of the random effects for the provider clusters. In this case, the distributional assumption about the random effects could be more critical to ensure consistent inference, even if the fixed effects are correctly specified. To assess if the random effects are normally distributed, a diagnostic test could be employed [22]. The impact of deviations from distributional assumptions for the random effects on the estimators in the presence of dissemination effect has not been fully studied, which directs us to conduct a simulation study.

## 5 Simulations

A simulation study was conducted to evaluate the performance of the direct, disseminated, composite, and overall effects estimators in scenarios with a binary outcome. We further evaluated the robustness of the Horvitz-Thompson and Hájek approaches when the normality assumption of the random effects in the treatment weight models was violated. We considered the impact of cluster size (equal or unequal) and random effects distribution (normal, left skew, right skew, or bimodal). The following quantities were computed for each scenario: the finite-sample bias in percent (%Bias), (estimates-truth)/truth for each estimate, empirical standard error (ESE), estimated average standard error (ASE), and empirical coverage probability (CP) of the 95% confidence intervals. A total of 1000 data sets were simulated per scenario as follows.

In this first scenario, we considered provider clusters of equal size. We considered *K* = 750 clusters with equal cluster size (*n*_*k*_ = 3 patients per cluster). We generated the following covariates based on the observed distribution in the motivating data set: binary age *X*_*i*1_ *∼* Bern(0.44); binary mean daily morphine milligram equivalence (MME) *X*_*i*2_ *∼* Bern(0.08); sex *X*_*i*3_ *∼* Bern(0.4); depression *X*_*i*4_ *∼* Bern(0.34); Charlson Comorbidity Index (CCI) *X*_*i*5_ followed the same observed distribution in the study data (mean = 1; sd = 1.15); and binary benzodiazepine use *X*_*i*6_ *∼* Bern(0.45). We generated the outcome as *Y*_*ki*_ *∼* Bern 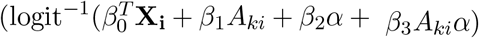 with *β* = (*β*_**0**_^*T*^, *β*_1_, *β*_2_, *β*_3_)^*T*^ = (*−*2.74, *−*1.00, 0.27, *−*0.13, 0.45, 0.09, 0.18, *−*0.50, *−*0.85, 0.88)^*T*^ and **X**_**i**_ = (1, *X*_*i*1_, …, *X*_*i*6_)^*T*^. The treatment *A*_*ki*_ *∼* Bern(logit^*−*1^(*γ*^*T*^ **X**_**i**_+*b*_*k*_)) with *γ* = (1.00, *−*0.41, *−*1.06, *−*0.09, *−*0.21, *−*0.10, 0.09)^*T*^. For the causal contrasts, we considered prescription coverage levels of 33%, 50%, and 67% in the provider cluster.

In our motivating example, the distribution of the random effects was left-skewed (Appendix Figure 1), which violates the normality assumption of the random effects for both estimators. To investigate the robustness of the two estimators under the normality assumption, we considered the following four scenarios when random effects follow a normal, a right-skewed generalized T, a left-skewed generalized T, and a symmetric bimodal distributions, separately for equal and unequal cluster sizes [58]. The complete simulation results for all scenarios are reported in the Appendix (Table A2 and Table A3).

Table 2 presents the simulation results when the random effects are left skewed with similar skewness of the random effects as in the motivating example with *equal* cluster sizes. We observed that the Hájek estimates performed better than the Horvitz-Thompson estimates, with Hájek estimates having smaller bias in percent and slightly smaller ESE and ASE. In addition, the ESEs were approximately equal to the ASEs, showing that the variance estimates employed from the inferference package [59] and the stabilizedinterference package [60] had reasonable finite sample performance in our setting. Even if the distribution of the random effects is not normal, there were little biases in the estimates with coverage probabilities around the nominal level. In Appendix Table A2, we reported the simulation results when the random effects follow a normal, a right-skewed, and a symmetric bimodal distributions, respectively. Results were robust under different distributions of random effects when we assumed equal cluster size.

**Table 2:**
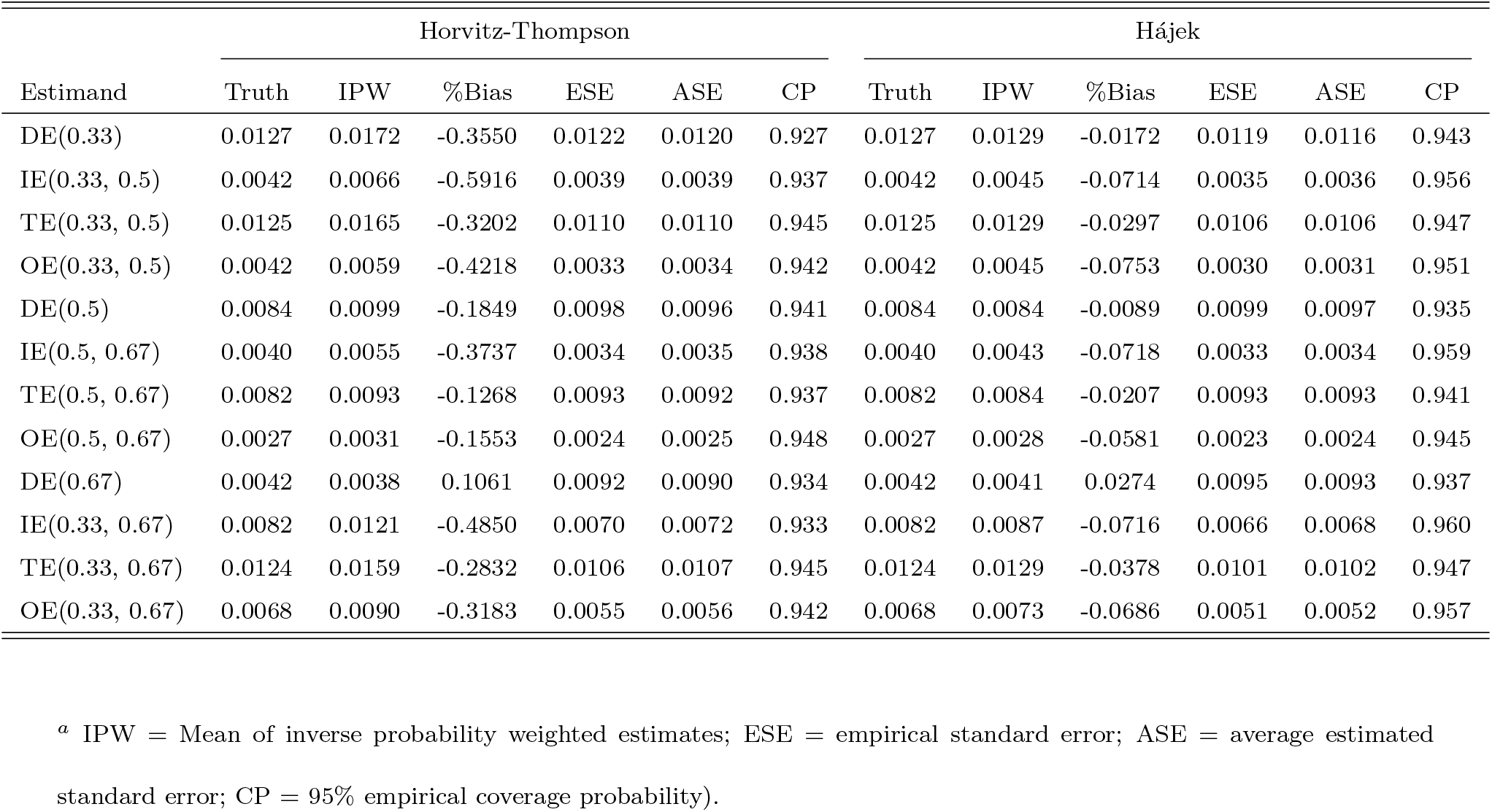
Simulation results with equal provider cluster sizes for direct, disseminated, composite and overall effects with left-skewed distribution for the random intercepts in the treatment weight model for 1000 simulated data sets.^*a*^

In this scenario with unequal cluster sizes, we considered *K* = 750 clusters with unequal sizes following the same empirical distribution (mean = 3.15) as in the motivating example. Other settings are the same as those in scenario with equal cluster sizes. Table 3 presents the simulation results with a left skewed distribution of random effects in treatment propensity score model. For unequal cluster size, we also considered the distribution of the random effects to be normal, left skew, right skew, and bimodal (Appendix Table A3). We first note that the Horvitz-Thompson and Hájek estimates performed well when the distribution of random effects is symmetric (normal and symmetric bimodal scenarios), but did not perform well when the distribution of the random effects is skewed (left-skewed and right-skewed scenarios). For example, some estimates were biased with large variance estimates and low coverage probabilities when the random effects were skewed to the left (Appendix Table A3). When the random effect distribution was skewed (left or right), the Hájek estimator performed better (in terms of empirical coverage probabilities) than the Horvitz-Thompson.

**Table 3:**
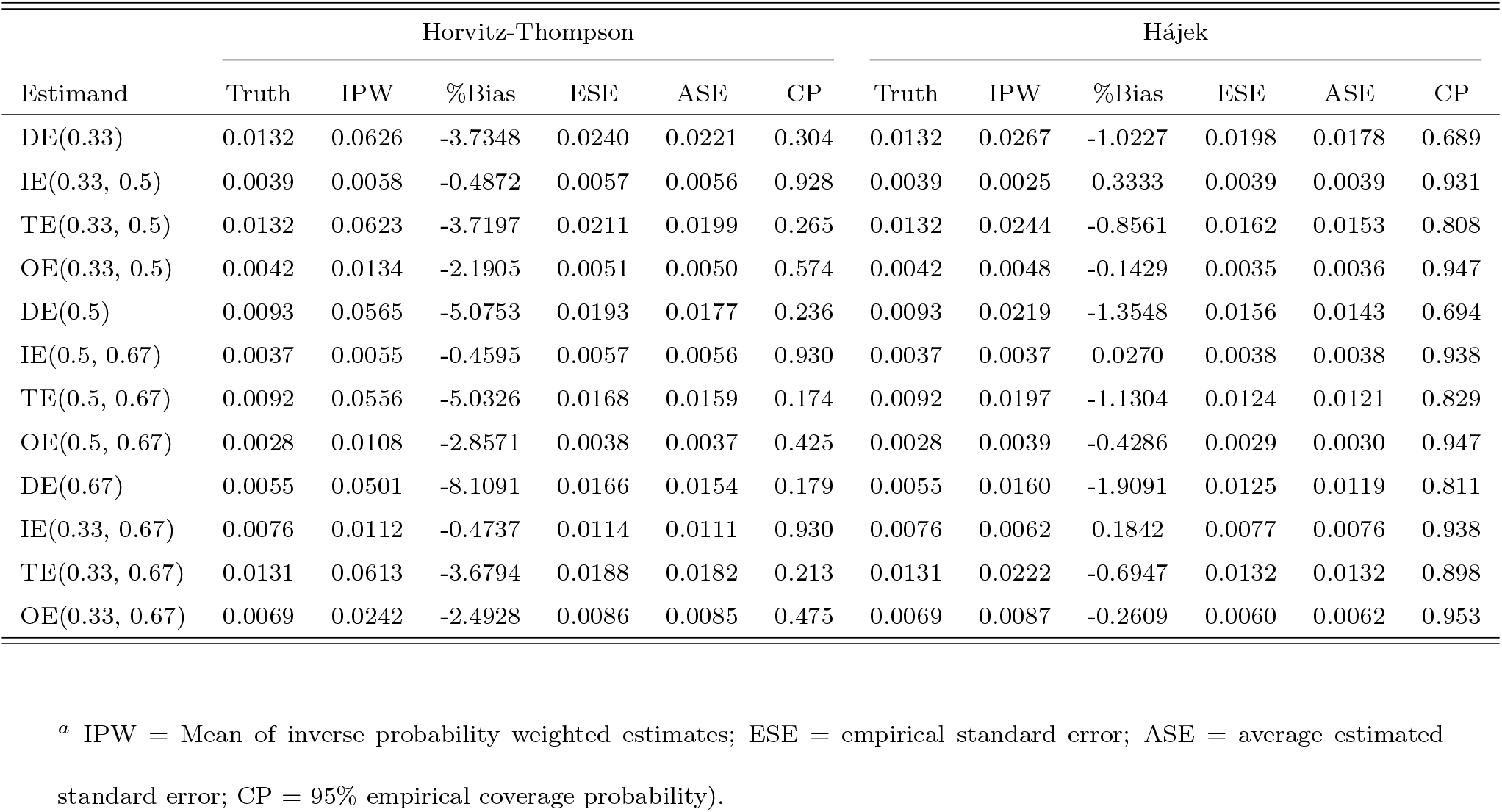
Simulation results with unequal provider cluster sizes for direct, disseminated, composite and overall effects with left-skewed distribution for the random intercepts in the treatment weight model for 1000 simulated data sets.^*a*^

These results of the simulation study suggest that the skewness of the random effect distributions may be leading to the worse performance of the Horvitz-Thompson estimator in this setting. Among the two approaches, the Hájek estimates tended to be less biased with smaller variances estimates and better coverage probabilities, suggesting that Hájek approach may be preferable with a skewed distribution of random effects in a study with the unequal cluster sizes. With equal cluster sizes, the Horvitz-Thompson and Hájek estimators both perform well in finite samples. Inference with equal cluster size tends to be more robust than unequal cluster size (which requires a larger effect size) under random effect misspecification [61]. As expected, the coverage probabilities in Table 3 are lower than those in Table 2 because the mixed effects model will lead to an increase in Type I error and a decrease in power with unequal cluster sizes when the random effects are misspecified [62, 63].

## 6 Application

### 6.1 Analysis

We employed a retrospective cohort study investigating the direct and disseminated effects of MOUD on the outcome opioid overdose among adults diagnosed with opioid use disorder (OUD). Our data was extracted from Optum’s de-identified Clinformatics® Data Mart Database, including patient eligibility, outpatient diagnosis, inpatient diagnosis, pharmacy visits and prescriptions from Jan 1, 2010 to Dec 31, 2017 [41]. The database used the International Classification of Diseases (ICD)-9 diagnosis before Sep 31, 2015 and switched to ICD-10 after Oct 1, 2015. The study timeline had 3 periods: baseline period, index period and follow-up. The baseline period was up to one year (i.e., 365 days) before the first diagnosis of OUD in the database which happened before Oct 1, 2015. The index period started with the first diagnosis of OUD and lasted for 90 days. The follow-up period began after the index period and ended at the time of the first opioid overdose, administrative censoring on Dec 31, 2017 or the patient left the database due to death or disenrollment, whichever came first (Figure 2). We did not allow for any gap in disenrollment (i.e., patient follow-up was censored at the time of disenrollment). The baseline period allowed for a pre-treatment period to ascertain information on possible confounders. The index period was primarily used to define the exposure window for MOUD initiation, following other published literature about the OUD treatment cascade and optimal timing for MOUD initiation following OUD diagnosis [64–66]. After an OUD diagnosis, providers are recommended to consider changing a patient’s opioid prescription due to their increased risk of overdose [67], as well as their prescriptions for benzodiazepines and other central nervous system depressants [68]. Using information about these often short acting prescription drugs ascertained during index period could potentially better capture a patient’s prescription status before the start of follow-up.

Framing this study as an ideal target trial can help to inform how we define the analytical sample and determine appropriate methodology for the analysis (Table 1). We included adults ages 18-90 years old who had been diagnosed with OUD and continuously enrolled in the database for both baseline and index period. Patients who had any buprenorphine-naloxone (BUP-NX) or injectable naltrexone prescription during the baseline period were excluded; patients who were diagnosed with opioid overdose during baseline period or index period were also excluded; patients who had received any methadone from a clinic during baseline period were excluded (as captured by Current Procedural Terminology (CPT) codes). The treatment strategy was defined as receipt of BUP-NX with at least 7 days of supply during the index period and remain on BUP-NX during follow-up, or refrain from taking BUP-NX during the index period and follow-up. In an ideal trial, provider clusters are first assigned to BUP-NX coverage levels, then patients are randomly assigned to either treatment strategy at baseline according to that coverage level. Follow-up starts at randomization and ends at the first opioid overdose, death, loss to follow-up, or two years after baseline, whichever occurs first. The outcome of interest is opioid overdose diagnosed by a medical professional within two years after baseline. The causal contrast of interest is the intention-to-treat effect estimated via comparison of two-year opioid overdose risks among patients assigned to each treatment strategy. Because treatment at both the provider and patient levels was not randomly assigned in this retrospective cohort study, we employed methods to adjust for baseline confounding. We adjusted for the following variables: age, gender, depression, CCI, mean daily morphine milligram equivalence (MME), and benzodiazepine prescription [69]. Depression and CCI were identified during baseline period, while MME and benzodiazepine prescription was identified during index period.

We developed an approach to build clusters among patients based on their main providers during the index period. We created an algorithm to identify the main provider using prescription claims. The first step was to identify each patient’s main provider. We considered claims with a sequence of BUP-NX prescriptions, other opioid prescriptions and all other prescriptions. Then, we selected the provider with the most prescriptions as the patient’s main provider. To break a tie between providers, we chose the most recently visited provider and then the provider who prescribed the highest MME. The second step was to determine which patients were included in each provider cluster using each patient’s main prescriber. National Provider Identifier (NPI) code was used to identify providers. If NPI was missing, Drug Enforcement Administration (DEA) code was used. Patients were clustered by their main provider. On average, each patient had three providers (range from 1 to 14 providers, standard deviation = 1.96) during the index period. After applying the algorithm to identify the main provider, there were 112 patients (5%) with a tie for their main provider. Among these 112 patients, patients had two tied providers on average (range from 2 to 5 providers). Then, we implemented our tie-breaking rule and selected the last provider after sorting by their NPI.

Clusters that had only one patient were excluded and the analysis included only provider clusters with at least some BUP-NX prescribing. We also required all patients in the same cluster to have visited the main provider within a two-year calendar window, where this window was defined from the the date of OUD diagnosis of the first patient for that particular provider in our study. We defined a two-level treatment: patient-level BUP-NX treatment was defined as BUP-NX prescription during the index period; cluster-level BUP-NX coverage was defined as the proportion of patients in the provider cluster who were prescribed BUP-NX during index period. Buprenorphine-naloxone was identified by brand and generic names. The outcome was the first diagnosis of opioid overdose during the follow-up period, as captured by ICD-9/ICD-10 diagnosis code in both inpatient and outpatient medical files. For the coverage levels selected in the analysis, we were interested in lower, moderate and higher coverage scenarios. Empirical studies suggest current MOUD coverage among those with OUD is approximately 20%, so we selected a scenario close to those empirical estimates to represent a lower coverage scenario [2, 70]. We also had to balance our decision with the observed distribution of coverage to ensure there were enough clusters in the database at (or around) each of the selected coverage levels, so we selected 33% coverage to represent lower, 50% to represent moderate, and 67% to represent higher coverage scenarios.

We included a complete list of the diagnosis codes in the Supplemental Appendix Table A4, as well as information on the validation and use of these codes. All database development and statistical analyses were performed with SAS version 9.4 (SAS Institute, Cary, NC) and R statistical software, version 3.2.3 (R Core Team 2016). For estimation of the disseminated effects, we employed an existing R package inferference [59] and the stabilizedinterference package (*v0*.*0*.*2*.*9200*) [60]. The study protocol was reviewed and approved by the University of Rhode Island Institutional Review Board.

### 6.2 Results

A total of 2,273 patients in 722 clusters were included and clusters included 3 patients on average and ranged in size from 2 to 19 patients, 64.3% were prescribed BUP-NX during the index period; 3.7% had nonfatal opioid overdose during follow-up; 40% were female; mean age was 37 years; and mean CCI was 1 (Table 4). During follow-up, 27 (1%) died. Patients were followed for a median of 587 days (Quartile 1 (Q1) = 232; Q3 = 730 days) and 1435 (63%) completed one year of follow-up and 1151 (51%) complted two years of follow-up. The insurance type included mostly commercial plans (88%) and some with Medicare Advantage plans (12%). The average daily MME was 60 mg in the provider clusters with higher MOUD coverage (*>*50%) and these provider practices had slightly younger patients on average, and less patients with Medicare Advantage plans, compared to lower coverage MOUD provider practices. Supplemental Table A1 displays the cumulative incidence of overdose overall by the end of the two year follow-up. The cumulative incidence was highest among provider clusters with lower coverage (*≤*33%). Among all patients, the cumulative incidence typically decreased as BUP-NX coverage increased. Based on a diagnostic test [22], we found evidence that the random effects for the treatment propensity score model were not normally distributed (*D* = 29.14, P value *<* 0.001). Based on a visual inspection, the distribution of the random effects was left skewed (Appendix Figure 1) and following our simulation results, this suggests that the Hájek-type estimators may be more appropriate in this particular study.

**Table 4:**
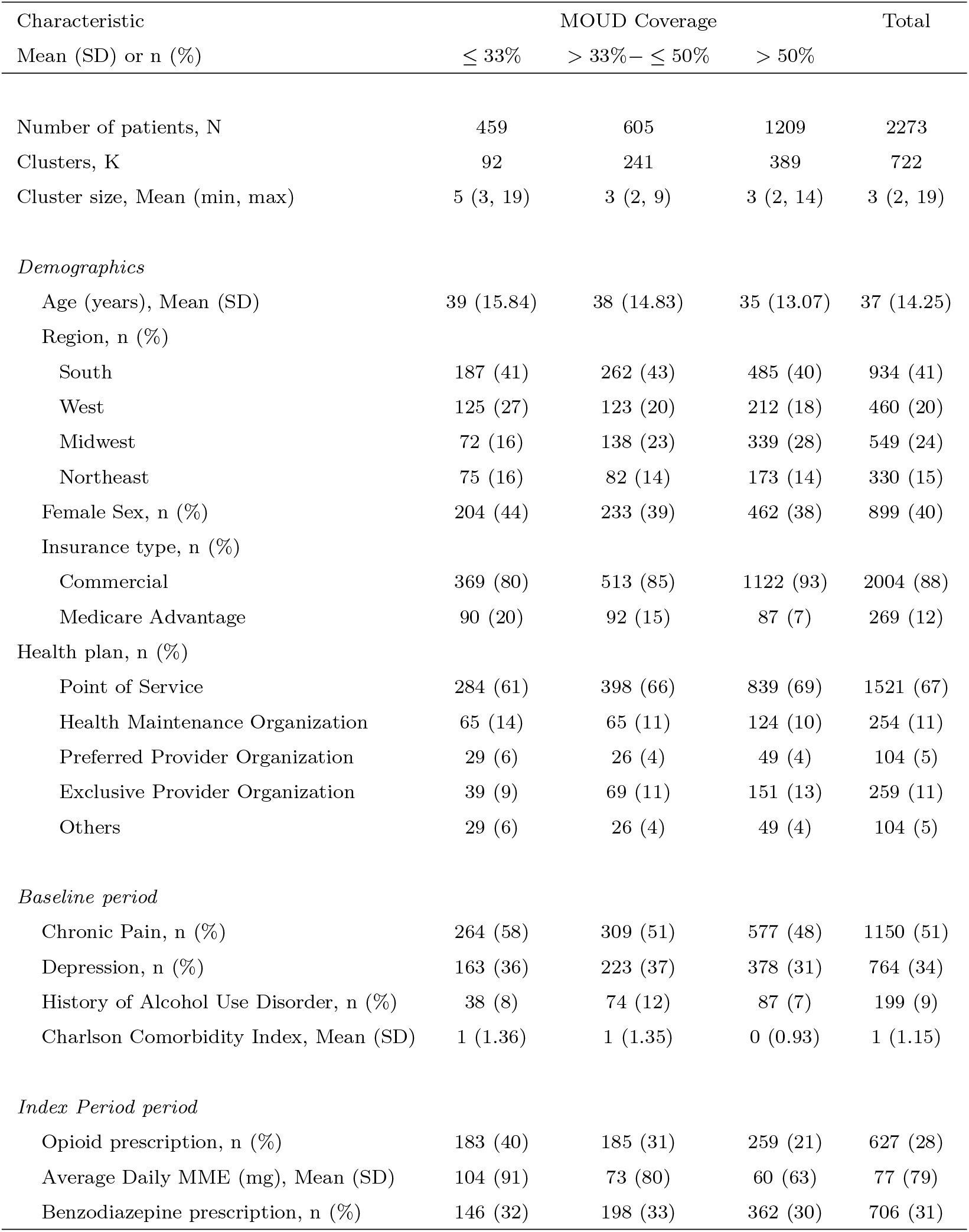
Characteristics of 2,273 patients diagnosed with opioid use disorder in Optum’s de-identified Clinformatics® Data Mart Database, 2010-2017, United States by medication for opioid use disorder (MOUD) coverage in provider clusters^*a*^

| Characteristic<br>Mean (SD) or n (%) | MOUD Coverage |  |  | Total |
| --- | --- | --- | --- | --- |
|  | ≤ 33% | > 33%– ≤ 50% | > 50% |  |
| Number of patients, N | 459 | 605 | 1209 | 2273 |
| Clusters, K | 92 | 241 | 389 | 722 |
| Cluster size, Mean (min, max) | 5 (3, 19) | 3 (2, 9) | 3 (2, 14) | 3 (2, 19) |
| *Demographics* |  |  |  |  |
| Age (years), Mean (SD) | 39 (15.84) | 38 (14.83) | 35 (13.07) | 37 (14.25) |
| Region, n (%) |  |  |  |  |
| South | 187 (41) | 262 (43) | 485 (40) | 934 (41) |
| West | 125 (27) | 123 (20) | 212 (18) | 460 (20) |
| Midwest | 72 (16) | 138 (23) | 339 (28) | 549 (24) |
| Northeast | 75 (16) | 82 (14) | 173 (14) | 330 (15) |
| Female Sex, n (%) | 204 (44) | 233 (39) | 462 (38) | 899 (40) |
| Insurance type, n (%) |  |  |  |  |
| Commercial | 369 (80) | 513 (85) | 1122 (93) | 2004 (88) |
| Medicare Advantage | 90 (20) | 92 (15) | 87 (7) | 269 (12) |
| Health plan, n (%) |  |  |  |  |
| Point of Service | 284 (61) | 398 (66) | 839 (69) | 1521 (67) |
| Health Maintenance Organization | 65 (14) | 65 (11) | 124 (10) | 254 (11) |
| Preferred Provider Organization | 29 (6) | 26 (4) | 49 (4) | 104 (5) |
| Exclusive Provider Organization | 39 (9) | 69 (11) | 151 (13) | 259 (11) |
| Others | 29 (6) | 26 (4) | 49 (4) | 104 (5) |
| *Baseline period* |  |  |  |  |
| Chronic Pain, n (%) | 264 (58) | 309 (51) | 577 (48) | 1150 (51) |
| Depression, n (%) | 163 (36) | 223 (37) | 378 (31) | 764 (34) |
| History of Alcohol Use Disorder, n (%) | 38 (8) | 74 (12) | 87 (7) | 199 (9) |
| Charlson Comorbidity Index, Mean (SD) | 1 (1.36) | 1 (1.35) | 0 (0.93) | 1 (1.15) |
| *Index Period period* |  |  |  |  |
| Opioid prescription, n (%) | 183 (40) | 185 (31) | 259 (21) | 627 (28) |
| Average Daily MME (mg), Mean (SD) | 104 (91) | 73 (80) | 60 (63) | 77 (79) |
| Benzodiazepine prescription, n (%) | 146 (32) | 198 (33) | 362 (30) | 706 (31) |

Table 5 displays the unadjusted and estimated risk differences of the causal effects of BUPNX on the likelihood of overdose using both the Horvitz-Thompson and Hájek-type estimators. Among clusters with no BUP-NX prescribing, the unadjusted cumulative incidence of opioid overdose was 2% (95% confidence interval (CI): 0.016, 0.021). We first describe the results obtained using the Horvitz-Thompson-type estimator, then the Hájek-type estimator. Adjusting for confounders age, gender, depression, CCI during the baseline period, benzodiazepine and MME during the index period and clustering by provider with a random intercept, we would expect 1 fewer overdose per 100 people (95% CI: -0.03, 0.01) for being treated in the high coverage (67%) provider clusters compared to being untreated in low coverage (33%) clusters. Furthermore, regardless of individual treatment status, there were an estimated 2 fewer overdoses per 100 people (95% CI: -0.03, -0.002) if 67% of the patients for a provider are treated compared to only if 33% are treated. For the estimated disseminated effect, we would expect 2 fewer overdoses per 100 people (95% CI: -0.04, -0.001) in untreated patients within high coverage clusters compared to within low coverage clusters. All direct effect estimates were close to the null when estimated using the Horvitz-Thompson-type estimator.

**Table 5:** Unadjusted and adjusted estimated risk differences (RDs) with corresponding 95% confidence intervals (CIs) of causal effects of medications for opioid use disorder (MOUD) on risk of overdose among 2,273 patients in Optum’s de-identified Clinformatics® Data Mart Database, 2010- 2017, United States.

| Effect | Coverage<br>($\alpha\%$, $a$) - ($\alpha'\%$, $a'$) | Unadjusted | | IPW Estimator | | Hájek Estimator | |
| --- | --- | --- | --- | --- | --- | --- | --- |
|  |  | RD | 95% CI | RD | 95% CI | RD | 95% CI |
| Direct | (33, 1) - (33, 0) | 0.012 | ( -0.011 , 0.035 ) | 0.001 | ( -0.028 , 0.03 ) | -0.028 | ( -0.056 , -0 ) |
| Direct | (50, 1) - (50, 1) | 0.009 | ( -0.009 , 0.027 ) | 0.01 | ( -0.009 , 0.029 ) | -0.008 | ( -0.029 , 0.013 ) |
| Direct | (67, 0) - (67, 1) | 0.006 | ( -0.012 , 0.023 ) | 0.01 | ( -0.009 , 0.029 ) | 0.002 | ( -0.016 , 0.019 ) |
| Disseminated | (50, 0) - (33, 0) | -0.004 | ( -0.012 , 0.004 ) | -0.016 | ( -0.031 , -0.002 ) | -0.017 | ( -0.031 , -0.002 ) |
| Disseminated | (67, 0) - (33, 0) | -0.004 | ( -0.016 , 0.008 ) | -0.02 | ( -0.038 , -0.001 ) | -0.029 | ( -0.05 , -0.008 ) |
| Disseminated | (67, 0) - (50, 0) | -0.001 | ( -0.005 , 0.004 ) | -0.003 | ( -0.009 , 0.002 ) | -0.012 | ( -0.021 , -0.003 ) |
| Composite | (50, 1) - (33, 0) | 0.005 | ( -0.013 , 0.024 ) | -0.006 | ( -0.031 , 0.018 ) | -0.013 | ( -0.024 , -0.002 ) |
| Composite | (67, 1) - (33, 0) | 0.002 | ( -0.015 , 0.018 ) | -0.012 | ( -0.034 , 0.011 ) | -0.02 | ( -0.035 , -0.005 ) |
| Composite | (67, 1) - (50, 0) | 0.005 | ( -0.011 , 0.021 ) | 0.005 | ( -0.012 , 0.022 ) | -0.007 | ( -0.013 , -0.002 ) |
| Overall | (50) - (33) | -0.003 | ( -0.009 , 0.003 ) | -0.012 | ( -0.022 , -0.001 ) | -0.025 | ( -0.049 , -0 ) |
| Overall | (67) - (33) | -0.004 | ( -0.013 , 0.004 ) | -0.015 | ( -0.028 , -0.002 ) | -0.027 | ( -0.051 , -0.004 ) |
| Overall | (67) - (50) | -0.001 | ( -0.005 , 0.003 ) | -0.003 | ( -0.007 , 0.002 ) | -0.011 | ( -0.03 , 0.008 ) |

The point estimates were somewhat further from the null when estimated using the Hájektype estimator, as compared to the Horvitz-Thompson-type estimators. For example, we would expect 2 fewer overdoses per 100 people (95% CI: -0.04, -0.01) for being treated in the high coverage (67%) provider clusters compared to being untreated in low (33%) coverage clusters. Regardless of individual treatment status, there were an estimated 3 fewer overdoses per 100 people (95% CI: -0.05, -0.00) if 67% of the patients for a provider are treated compared to only if 33% are treated. For the estimated disseminated effect, we would expect 3 fewer overdoses per 100 people (95% CI: -0.05, -0.01) in untreated patients within high coverage clusters compared to within low coverage clusters. The direct effect estimate with the Hajèk estimator was protective for the 33% coverage clusters (95% CI: -0.06, -0.00) with an estimated 3 fewer cases per 100 people for being treated versus untreated within 33% coverage clusters.

## 7 Discussion

We presented a novel application of causal inference methods to evaluate dissemination of baseline prescription for medications for opioid use disorder to prevent overdose among a cohort of patients with diagnosed opioid use disorder and adjusted for baseline confounders using information from an administrative claims database. For the estimated composite effect, we would expect 2 fewer opioid overdoses per 100 people in treated patients within high coverage provider clusters compared to untreated patients within low coverage clusters. For the estimated disseminated effect, we would expect 3 fewer overdoses per 100 people in untreated patients within high coverage clusters compared to within low coverage clusters. This study provides preliminary evidence that, in addition to increasing the number of MOUD prescribers, increasing MOUD coverage within prescriber practices could offer additional benefits among patients with opioid use disorder. These results suggest possible medication diversion; however, the methods are agnostic to the spillover mechanism. The observed reductions in overdose for the disseminated effect may be due to spillover from medication diversion, shared provider norms, geographic proximity, or another mechanism and are prone to biases common in administrative data, such as measurement error of the outcomes. Future primary data collection studies could investigate medication diversion by collecting information on self-reported MOUD from sources other than medical providers [38, 39] and use this information to further disentangle spillover benefits of MOUD.

In our simulation study, the Hájek estimator had better finite sample performance for both the left and right skew distribution of the random effects when there were unequal cluster sizes. The Hájek (stabilized) estimator employs an unbiased estimator of the cluster size, instead of the observed cluster size. If the propensity score is very small (or very large), the numerator essentially tracks the denominator, which can reduce variability in the estimator. In Liu, et al. [56], Hájek estimators were biased if the fixed effects were mis-specified; however, their second Hájek estimator may be more robust to misspecification of the propensity score than their Horvitz-Thompson estimator or their first Hájek estimator. Herein, we employed the second Hájek estimator from Liu, et al. (2016) [56]. Intuitively, if the propensity score model is incorrect and this term is included in both the numerator and denominator, the impact of that misspecification on estimation of the causal parameter may be reduced.

Studies that analyze administrative claims databases have specific features that cannot be fully addressed by commonly-used methods for prospective cohort study designs and an important shortcoming of commonly used methods for dissemination is that they do not take into account dissemination between provider clusters. We suspect in this analysis that we may have unmeasured confounding because some of the estimated direct effects were close to the null, possibly due to unmeasured disease severity or lifestyle factors like illicit drug use. There have been limited methods developments for test negative designs [71], negative controls, [72] or interrupted time series [73] with dissemination. We recommend this as an area for future work, particularly with applications to routinely-collected health data. We acknowledge that MOUD prescriptions are a time-varying exposure. Although the follow-up was limited to two years, patient exposure to MOUD could have changed after the index period. In future work, a time-varying exposure and corresponding time-varying interference set, as well as different versions of the exposure, could be considered. We assumed that dissemination occurs within, but not between provider-based clusters; however, many providers work in group practice settings. There could be some degree of influence between patients who are treated in group practice settings, yet this was not measurable in our data. If we had more complete network information, we would have the opportunity to consider different exposure mappings based on position and influence in the network. Absent that information, we assume that it does not matter who was treated, but rather how many were treated in a cluster. When cluster sizes vary largely, this assumption can be dubious. In our setting, the cluster sizes ranged from 2 to 19 patients, so this may be a strong assumption particularly for the larger clusters.

Statistical analyses in administrative health claims databases face additional considerations, such as misclassification, informative drop out, and unmeasured confounding. In this study, we did not evaluate receipt of prescription naloxone because this is likely not well captured in administrative claims due to distribution and administration in the community. Although prescription information from a pharmacy is considered to be accurate, misclassification of the patient’s duration of exposure can occur, for example, if a patient decides to extend the prescription by taking a lower dose. It is also possible that patients sought treatment at clinics (e.g., methadone) that did not submit claims for the care received by the patient. Furthermore, some overdoses may not be captured in the administrative claims data, specifically those treated in the exclusively in the community typically with naloxone or those who died before reaching the hospital. For the definition of OUD, we used sets of claims from the literature [66, 74]; however, our definition is not a validated measure and does not capture disease severity. Standard estimation methods assume that there is no bias due study drop out. If this assumption is questionable, censoring weights could be employed in the analysis; however, selection bias in routinely-collected health data may complicate application of existing methods due to different censoring mechanisms [75, 76].

Based on our simulation study, we recommend extending these estimators for alternative distributions of the random effects to better match distributions observed in the data [62, 63, 77], as well as possibly a generalized estimating equation (GEE) approach to quantify the cluster-level exposure weights [78]. A GEE model would be a way to avoid the issue with the normality of the random effects observed with linear mixed models in this setting; however, this approach would make different assumptions about modeling the cluster-level treatment. In future work, we will extend our methods allowing for opioid prescription levels to be time varying by evaluating the average coverage level within a predefined time window (e.g., one calendar month) and individual patients’ observed opioid prescription history. Studies that leverage information from administrative claims offer the possibility of better informing MOUD prescribing practices at the provider level to provide a maximal reduction in overdoses among people with opioid use disorder.

## Data Availability

Restrictions apply to the availability of Optum's de-identified Clinformatics Data Mart Database. Data was obtained from Optum through a third-party license and authors cannot make these data publicly available due to the data use agreement.

## ABBREVIATIONS

(BUP-NX): Buprenorphine-naloxone
(CPT): Current Procedural Terminology
(DEA): Drug Enforcement Administration
(ESE): Empirical standard error
(ASE): Estimated average standard error
(ECP): Empirical coverage probability
(ICD): International classification of diseases
(MOUD): Medications for opioid use disorder
(NPI): National Provider Identifier
(OUD): Opioid use disorder

