## Supplemental File for "Toward Evaluation of Disseminated Effects of Medications for Opioid Use Disorder within Provider-Based Clusters Using Routinely-Collected Health Data"

for

April 12, 2022

### Appendix: Supplemental Tables and Figures

Table A1: Cumulative incidence of overdose in Optum's de-identified Clinformatics® Data Mart Database, 2010-2015, United States

| | All patients | | | |
|---|---|---|---|---|
| MOUD Coverage | Total Person | Overdose | Cum Inc (%) | (95%CI) |
| $\leq 33\%$ | 459 | 19 | 4.14 | (2.3, 5.9) |
| $> 33\% - \leq 50\%$ | 605 | 25 | 4.13 | (2.5, 5.7) |
| $> 50\%$ | 1,209 | 39 | 3.2 | (2.2, 4.2) |
| All | 2,273 | 83 | 3.7 | (2.9, 4.5) |

Table A2: Simulation results with equal cluster sizes for direct, disseminated, composite and overall effects with different distributions for the random intercepts in the cluster-level propensity score model for 1000 simulated data sets.[a]

| | Horvitz-Thompson | | | | | | Hájek | | | | | |
|---|---|---|---|---|---|---|---|---|---|---|---|---|
| Estimand | Truth | IPW | %Bias | ESE | ASE | CP | Truth | IPW | %Bias | ESE | ASE | CP |
| Normal | | | | | | | | | | | | |
| DE(0.33) | 0.0127 | 0.0164 | -0.2916 | 0.0122 | 0.0117 | 0.925 | 0.0127 | 0.0128 | -0.0107 | 0.0119 | 0.0118 | 0.942 |
| IE(0.33, 0.5) | 0.0042 | 0.0057 | -0.3703 | 0.0040 | 0.0038 | 0.927 | 0.0042 | 0.0040 | 0.0449 | 0.0036 | 0.0036 | 0.930 |
| TE(0.33, 0.5) | 0.0125 | 0.0159 | -0.2657 | 0.0114 | 0.0108 | 0.926 | 0.0125 | 0.0126 | -0.0019 | 0.0110 | 0.0107 | 0.941 |
| OE(0.33, 0.5) | 0.0042 | 0.0054 | -0.2928 | 0.0035 | 0.0033 | 0.940 | 0.0042 | 0.0040 | 0.0290 | 0.0031 | 0.0030 | 0.931 |
| DE(0.5) | 0.0084 | 0.0101 | -0.2136 | 0.0100 | 0.0095 | 0.928 | 0.0084 | 0.0086 | -0.0252 | 0.0101 | 0.0099 | 0.941 |
| IE(0.5, 0.67) | 0.0040 | 0.0051 | -0.2818 | 0.0036 | 0.0034 | 0.925 | 0.0040 | 0.0038 | 0.0469 | 0.0035 | 0.0033 | 0.941 |
| TE(0.5, 0.67) | 0.0082 | 0.0094 | -0.1395 | 0.0097 | 0.0092 | 0.941 | 0.0082 | 0.0083 | -0.0116 | 0.0097 | 0.0094 | 0.942 |
| OE(0.5, 0.67) | 0.0027 | 0.0029 | -0.0909 | 0.0026 | 0.0025 | 0.944 | 0.0027 | 0.0025 | 0.0409 | 0.0025 | 0.0024 | 0.944 |
| DE(0.67) | 0.0042 | 0.0043 | -0.0054 | 0.0094 | 0.0090 | 0.945 | 0.0042 | 0.0045 | -0.0667 | 0.0097 | 0.0095 | 0.946 |
| IE(0.33, 0.67) | 0.0082 | 0.0108 | -0.3270 | 0.0073 | 0.0070 | 0.925 | 0.0082 | 0.0078 | 0.0459 | 0.0069 | 0.0066 | 0.938 |
| TE(0.33, 0.67) | 0.0124 | 0.0151 | -0.2172 | 0.0111 | 0.0105 | 0.934 | 0.0124 | 0.0123 | 0.0074 | 0.0106 | 0.0102 | 0.933 |
| OE(0.33, 0.67) | 0.0068 | 0.0083 | -0.2143 | 0.0058 | 0.0055 | 0.940 | 0.0068 | 0.0066 | 0.0336 | 0.0054 | 0.0052 | 0.944 |
| Right Skewed | | | | | | | | | | | | |
| DE(0.33) | 0.0127 | 0.0153 | -0.2090 | 0.0117 | 0.0116 | 0.941 | 0.0127 | 0.0124 | 0.0181 | 0.0114 | 0.0113 | 0.940 |
| IE(0.33, 0.5) | 0.0042 | 0.0055 | -0.3159 | 0.0039 | 0.0037 | 0.934 | 0.0042 | 0.0042 | -0.0079 | 0.0036 | 0.0034 | 0.938 |
| TE(0.33, 0.5) | 0.0125 | 0.0152 | -0.2168 | 0.0107 | 0.0107 | 0.948 | 0.0125 | 0.0123 | 0.0172 | 0.0103 | 0.0104 | 0.947 |
| OE(0.33, 0.5) | 0.0042 | 0.0053 | -0.2748 | 0.0033 | 0.0032 | 0.952 | 0.0042 | 0.0041 | 0.0134 | 0.0030 | 0.0030 | 0.935 |
| DE(0.5) | 0.0084 | 0.0098 | -0.1675 | 0.0095 | 0.0094 | 0.946 | 0.0084 | 0.0081 | 0.0297 | 0.0095 | 0.0095 | 0.940 |
| IE(0.5, 0.67) | 0.0040 | 0.0053 | -0.3314 | 0.0034 | 0.0033 | 0.919 | 0.0040 | 0.0039 | 0.0109 | 0.0033 | 0.0032 | 0.938 |
| TE(0.5, 0.67) | 0.0082 | 0.0091 | -0.1122 | 0.0091 | 0.0091 | 0.947 | 0.0082 | 0.0080 | 0.0292 | 0.0091 | 0.0091 | 0.939 |
| OE(0.5, 0.67) | 0.0027 | 0.0030 | -0.1314 | 0.0024 | 0.0025 | 0.954 | 0.0027 | 0.0026 | 0.0176 | 0.0023 | 0.0024 | 0.952 |
| DE(0.67) | 0.0042 | 0.0038 | 0.0946 | 0.0091 | 0.0089 | 0.937 | 0.0042 | 0.0040 | 0.0465 | 0.0093 | 0.0091 | 0.938 |
| IE(0.33, 0.67) | 0.0082 | 0.0108 | -0.3234 | 0.0071 | 0.0068 | 0.933 | 0.0082 | 0.0081 | 0.0013 | 0.0067 | 0.0064 | 0.944 |
| TE(0.33, 0.67) | 0.0124 | 0.0146 | -0.1807 | 0.0104 | 0.0104 | 0.950 | 0.0124 | 0.0122 | 0.0167 | 0.0099 | 0.0100 | 0.943 |
| OE(0.33, 0.67) | 0.0068 | 0.0083 | -0.2191 | 0.0054 | 0.0055 | 0.955 | 0.0068 | 0.0067 | 0.0150 | 0.0050 | 0.0051 | 0.946 |
| Left Skewed | | | | | | | | | | | | |
| DE(0.33) | 0.0127 | 0.0172 | -0.3550 | 0.0122 | 0.0120 | 0.927 | 0.0127 | 0.0129 | -0.0172 | 0.0119 | 0.0116 | 0.943 |
| IE(0.33, 0.5) | 0.0042 | 0.0066 | -0.5916 | 0.0039 | 0.0039 | 0.937 | 0.0042 | 0.0045 | -0.0714 | 0.0035 | 0.0036 | 0.956 |
| TE(0.33, 0.5) | 0.0125 | 0.0165 | -0.3202 | 0.0110 | 0.0110 | 0.945 | 0.0125 | 0.0129 | -0.0297 | 0.0106 | 0.0106 | 0.947 |
| OE(0.33, 0.5) | 0.0042 | 0.0059 | -0.4218 | 0.0033 | 0.0034 | 0.942 | 0.0042 | 0.0045 | -0.0753 | 0.0030 | 0.0031 | 0.951 |
| DE(0.5) | 0.0084 | 0.0099 | -0.1849 | 0.0098 | 0.0096 | 0.941 | 0.0084 | 0.0084 | -0.0089 | 0.0099 | 0.0097 | 0.935 |
| IE(0.5, 0.67) | 0.0040 | 0.0055 | -0.3737 | 0.0034 | 0.0035 | 0.938 | 0.0040 | 0.0043 | -0.0718 | 0.0033 | 0.0034 | 0.959 |
| TE(0.5, 0.67) | 0.0082 | 0.0093 | -0.1268 | 0.0093 | 0.0092 | 0.937 | 0.0082 | 0.0084 | -0.0207 | 0.0093 | 0.0093 | 0.941 |
| OE(0.5, 0.67) | 0.0027 | 0.0031 | -0.1553 | 0.0024 | 0.0025 | 0.948 | 0.0027 | 0.0028 | -0.0581 | 0.0023 | 0.0024 | 0.945 |
| DE(0.67) | 0.0042 | 0.0038 | 0.1061 | 0.0092 | 0.0090 | 0.934 | 0.0042 | 0.0041 | 0.0274 | 0.0095 | 0.0093 | 0.937 |
| IE(0.33, 0.67) | 0.0082 | 0.0121 | -0.4850 | 0.0070 | 0.0072 | 0.933 | 0.0082 | 0.0087 | -0.0716 | 0.0066 | 0.0068 | 0.960 |
| TE(0.33, 0.67) | 0.0124 | 0.0159 | -0.2832 | 0.0106 | 0.0107 | 0.945 | 0.0124 | 0.0129 | -0.0378 | 0.0101 | 0.0102 | 0.947 |
| OE(0.33, 0.67) | 0.0068 | 0.0090 | -0.3183 | 0.0055 | 0.0056 | 0.942 | 0.0068 | 0.0073 | -0.0686 | 0.0051 | 0.0052 | 0.957 |
| Bimodal | | | | | | | | | | | | |
| DE(0.33) | 0.0127 | 0.0158 | -0.2493 | 0.0121 | 0.0116 | 0.936 | 0.0127 | 0.0126 | 0.0032 | 0.0118 | 0.0114 | 0.946 |
| IE(0.33, 0.5) | 0.0042 | 0.0054 | -0.2975 | 0.0037 | 0.0037 | 0.939 | 0.0042 | 0.0039 | 0.0540 | 0.0034 | 0.0034 | 0.940 |
| TE(0.33, 0.5) | 0.0125 | 0.0157 | -0.2505 | 0.0111 | 0.0106 | 0.937 | 0.0125 | 0.0125 | 0.0050 | 0.0108 | 0.0104 | 0.936 |
| OE(0.33, 0.5) | 0.0042 | 0.0053 | -0.2758 | 0.0033 | 0.0032 | 0.940 | 0.0042 | 0.0040 | 0.0381 | 0.0030 | 0.0030 | 0.924 |
| DE(0.5) | 0.0084 | 0.0103 | -0.2271 | 0.0100 | 0.0095 | 0.938 | 0.0084 | 0.0085 | -0.0194 | 0.0100 | 0.0096 | 0.934 |
| IE(0.5, 0.67) | 0.0040 | 0.0052 | -0.2987 | 0.0033 | 0.0033 | 0.937 | 0.0040 | 0.0038 | 0.0533 | 0.0032 | 0.0032 | 0.945 |
| TE(0.5, 0.67) | 0.0082 | 0.0096 | -0.1652 | 0.0097 | 0.0091 | 0.938 | 0.0082 | 0.0083 | -0.0104 | 0.0097 | 0.0092 | 0.934 |
| OE(0.5, 0.67) | 0.0027 | 0.0030 | -0.1313 | 0.0026 | 0.0025 | 0.932 | 0.0027 | 0.0026 | 0.0341 | 0.0025 | 0.0024 | 0.930 |
| DE(0.67) | 0.0042 | 0.0044 | -0.0393 | 0.0095 | 0.0090 | 0.938 | 0.0042 | 0.0045 | -0.0705 | 0.0097 | 0.0093 | 0.933 |
| IE(0.33, 0.67) | 0.0082 | 0.0106 | -0.2981 | 0.0068 | 0.0067 | 0.940 | 0.0082 | 0.0077 | 0.0536 | 0.0064 | 0.0064 | 0.944 |
| TE(0.33, 0.67) | 0.0124 | 0.0150 | -0.2097 | 0.0109 | 0.0103 | 0.934 | 0.0124 | 0.0123 | 0.0113 | 0.0104 | 0.0100 | 0.933 |
| OE(0.33, 0.67) | 0.0068 | 0.0083 | -0.2197 | 0.0056 | 0.0054 | 0.938 | 0.0068 | 0.0066 | 0.0365 | 0.0052 | 0.0051 | 0.937 |

[a] IPW = Mean of inverse probability weighted estimates; ESE = empirical standard error; ASE = average estimated standard error; CP = empirical coverage probability).

Table A3: Simulation results with unequal cluster sizes for direct, disseminated, composite and overall effects with different distributions for the random intercepts in the cluster-level propensity score model for 1000 simulated data sets.[a]

| | Horvitz-Thompson | | | | | | Hájek | | | | | |
|---|---|---|---|---|---|---|---|---|---|---|---|---|
| Estimand | Truth | IPW | %Bias | ESE | ASE | CP | Truth | IPW | %Bias | ESE | ASE | CP |
| Normal | | | | | | | | | | | | |
| DE(0.33) | 0.0132 | 0.0165 | -0.2500 | 0.0123 | 0.0122 | 0.939 | 0.0132 | 0.0116 | 0.1212 | 0.0124 | 0.0134 | 0.945 |
| IE(0.33, 0.5) | 0.0039 | 0.0051 | -0.3333 | 0.0036 | 0.0036 | 0.948 | 0.0039 | 0.0042 | -0.0769 | 0.0045 | 0.0053 | 0.959 |
| TE(0.33, 0.5) | 0.0132 | 0.0163 | -0.2424 | 0.0115 | 0.0114 | 0.943 | 0.0132 | 0.0116 | 0.1212 | 0.0111 | 0.0120 | 0.930 |
| OE(0.33, 0.5) | 0.0042 | 0.0053 | -0.2619 | 0.0033 | 0.0032 | 0.935 | 0.0042 | 0.0040 | 0.0238 | 0.0038 | 0.0044 | 0.932 |
| DE(0.5) | 0.0093 | 0.0112 | -0.2043 | 0.0105 | 0.0103 | 0.938 | 0.0093 | 0.0074 | 0.2043 | 0.0099 | 0.0102 | 0.930 |
| IE(0.5, 0.67) | 0.0037 | 0.0046 | -0.2162 | 0.0034 | 0.0033 | 0.929 | 0.0037 | 0.0039 | -0.0270 | 0.0043 | 0.0043 | 0.934 |
| TE(0.5, 0.67) | 0.0092 | 0.0107 | -0.1522 | 0.0101 | 0.0100 | 0.937 | 0.0092 | 0.0075 | 0.1848 | 0.0095 | 0.0097 | 0.929 |
| OE(0.5, 0.67) | 0.0028 | 0.0031 | -0.1071 | 0.0026 | 0.0025 | 0.948 | 0.0028 | 0.0026 | 0.0357 | 0.0031 | 0.0030 | 0.927 |
| DE(0.67) | 0.0055 | 0.0061 | -0.1091 | 0.0099 | 0.0098 | 0.940 | 0.0055 | 0.0037 | 0.3455 | 0.0093 | 0.0095 | 0.930 |
| IE(0.33, 0.67) | 0.0076 | 0.0097 | -0.2763 | 0.0068 | 0.0067 | 0.931 | 0.0076 | 0.0081 | -0.0658 | 0.0079 | 0.0088 | 0.947 |
| TE(0.33, 0.67) | 0.0131 | 0.0158 | -0.2061 | 0.0112 | 0.0111 | 0.940 | 0.0131 | 0.0117 | 0.1069 | 0.0106 | 0.0115 | 0.928 |
| OE(0.33, 0.67) | 0.0069 | 0.0083 | -0.2029 | 0.0056 | 0.0055 | 0.947 | 0.0069 | 0.0067 | 0.0290 | 0.0061 | 0.0067 | 0.922 |
| Right Skewed | | | | | | | | | | | | |
| DE(0.33) | 0.0132 | 0.0589 | -3.4545 | 0.0178 | 0.0172 | 0.271 | 0.0132 | 0.0347 | -1.6212 | 0.0155 | 0.0160 | 0.585 |
| IE(0.33, 0.5) | 0.0039 | 0.0033 | 0.1282 | 0.0044 | 0.0045 | 0.943 | 0.0039 | 0.0038 | 0.0256 | 0.0036 | 0.0040 | 0.955 |
| TE(0.33, 0.5) | 0.0132 | 0.0592 | -3.4848 | 0.0159 | 0.0155 | 0.178 | 0.0132 | 0.0327 | -1.4773 | 0.0133 | 0.0140 | 0.602 |
| OE(0.33, 0.5) | 0.0042 | 0.0118 | -1.8333 | 0.0041 | 0.0041 | 0.536 | 0.0042 | 0.0064 | -0.5476 | 0.0033 | 0.0035 | 0.916 |
| DE(0.5) | 0.0093 | 0.0559 | -5.0108 | 0.0142 | 0.0137 | 0.132 | 0.0093 | 0.0289 | -2.1075 | 0.0121 | 0.0124 | 0.527 |
| IE(0.5, 0.67) | 0.0037 | 0.0033 | 0.1081 | 0.0044 | 0.0045 | 0.945 | 0.0037 | 0.0046 | -0.2432 | 0.0034 | 0.0034 | 0.953 |
| TE(0.5, 0.67) | 0.0092 | 0.0556 | -5.0326 | 0.0126 | 0.0123 | 0.049 | 0.0092 | 0.0266 | -1.8913 | 0.0101 | 0.0108 | 0.564 |
| OE(0.5, 0.67) | 0.0028 | 0.0104 | -2.7143 | 0.0031 | 0.0031 | 0.267 | 0.0028 | 0.0051 | -0.8214 | 0.0027 | 0.0028 | 0.886 |
| DE(0.67) | 0.0055 | 0.0522 | -8.4909 | 0.0121 | 0.0117 | 0.037 | 0.0055 | 0.0220 | -3.0000 | 0.0097 | 0.0103 | 0.558 |
| IE(0.33, 0.67) | 0.0076 | 0.0067 | 0.1184 | 0.0088 | 0.0089 | 0.943 | 0.0076 | 0.0084 | -0.1053 | 0.0069 | 0.0074 | 0.956 |
| TE(0.33, 0.67) | 0.0131 | 0.0589 | -3.4962 | 0.0145 | 0.0143 | 0.097 | 0.0131 | 0.0304 | -1.3206 | 0.0116 | 0.0126 | 0.666 |
| OE(0.33, 0.67) | 0.0069 | 0.0222 | -2.2174 | 0.0070 | 0.0069 | 0.399 | 0.0069 | 0.0115 | -0.6667 | 0.0057 | 0.0060 | 0.904 |
| Left Skewed | | | | | | | | | | | | |
| DE(0.33) | 0.0132 | 0.0626 | -3.7348 | 0.0240 | 0.0221 | 0.304 | 0.0132 | 0.0267 | -1.0227 | 0.0198 | 0.0178 | 0.689 |
| IE(0.33, 0.5) | 0.0039 | 0.0058 | -0.4872 | 0.0057 | 0.0056 | 0.928 | 0.0039 | 0.0025 | 0.3333 | 0.0039 | 0.0039 | 0.931 |
| TE(0.33, 0.5) | 0.0132 | 0.0623 | -3.7197 | 0.0211 | 0.0199 | 0.265 | 0.0132 | 0.0244 | -0.8561 | 0.0162 | 0.0153 | 0.808 |
| OE(0.33, 0.5) | 0.0042 | 0.0134 | -2.1905 | 0.0051 | 0.0050 | 0.574 | 0.0042 | 0.0048 | -0.1429 | 0.0035 | 0.0036 | 0.947 |
| DE(0.5) | 0.0093 | 0.0565 | -5.0753 | 0.0193 | 0.0177 | 0.236 | 0.0093 | 0.0219 | -1.3548 | 0.0156 | 0.0143 | 0.694 |
| IE(0.5, 0.67) | 0.0037 | 0.0055 | -0.4595 | 0.0057 | 0.0056 | 0.930 | 0.0037 | 0.0037 | 0.0270 | 0.0038 | 0.0038 | 0.938 |
| TE(0.5, 0.67) | 0.0092 | 0.0556 | -5.0326 | 0.0168 | 0.0159 | 0.174 | 0.0092 | 0.0197 | -1.1304 | 0.0124 | 0.0121 | 0.829 |
| OE(0.5, 0.67) | 0.0028 | 0.0108 | -2.8571 | 0.0038 | 0.0037 | 0.425 | 0.0028 | 0.0039 | -0.4286 | 0.0029 | 0.0030 | 0.947 |
| DE(0.67) | 0.0055 | 0.0501 | -8.1091 | 0.0166 | 0.0154 | 0.179 | 0.0055 | 0.0160 | -1.9091 | 0.0125 | 0.0119 | 0.811 |
| IE(0.33, 0.67) | 0.0076 | 0.0112 | -0.4737 | 0.0114 | 0.0111 | 0.930 | 0.0076 | 0.0062 | 0.1842 | 0.0077 | 0.0076 | 0.938 |
| TE(0.33, 0.67) | 0.0131 | 0.0613 | -3.6794 | 0.0188 | 0.0182 | 0.213 | 0.0131 | 0.0222 | -0.6947 | 0.0132 | 0.0132 | 0.898 |
| OE(0.33, 0.67) | 0.0069 | 0.0242 | -2.4928 | 0.0086 | 0.0085 | 0.475 | 0.0069 | 0.0087 | -0.2609 | 0.0060 | 0.0062 | 0.953 |
| Bimodal | | | | | | | | | | | | |
| DE(0.33) | 0.0132 | 0.0161 | -0.2121 | 0.0126 | 0.0122 | 0.941 | 0.0132 | 0.0114 | 0.1439 | 0.0109 | 0.0108 | 0.944 |
| IE(0.33, 0.5) | 0.0039 | 0.0049 | -0.2821 | 0.0036 | 0.0035 | 0.928 | 0.0039 | 0.0036 | 0.0513 | 0.0040 | 0.0037 | 0.944 |
| TE(0.33, 0.5) | 0.0132 | 0.0163 | -0.2424 | 0.0116 | 0.0113 | 0.944 | 0.0132 | 0.0113 | 0.1364 | 0.0098 | 0.0098 | 0.943 |
| OE(0.33, 0.5) | 0.0042 | 0.0053 | -0.2857 | 0.0033 | 0.0032 | 0.947 | 0.0042 | 0.0037 | 0.1190 | 0.0032 | 0.0031 | 0.925 |
| DE(0.5) | 0.0093 | 0.0114 | -0.2258 | 0.0104 | 0.0103 | 0.946 | 0.0093 | 0.0077 | 0.1720 | 0.0094 | 0.0093 | 0.939 |
| IE(0.5, 0.67) | 0.0037 | 0.0049 | -0.3243 | 0.0035 | 0.0033 | 0.924 | 0.0037 | 0.0035 | 0.0541 | 0.0043 | 0.0039 | 0.940 |
| TE(0.5, 0.67) | 0.0092 | 0.0110 | -0.1957 | 0.0100 | 0.0100 | 0.944 | 0.0092 | 0.0077 | 0.1630 | 0.0092 | 0.0090 | 0.935 |
| OE(0.5, 0.67) | 0.0028 | 0.0033 | -0.1786 | 0.0025 | 0.0025 | 0.947 | 0.0028 | 0.0025 | 0.0714 | 0.0031 | 0.0029 | 0.932 |
| DE(0.67) | 0.0055 | 0.0061 | -0.1091 | 0.0099 | 0.0098 | 0.952 | 0.0055 | 0.0042 | 0.2364 | 0.0098 | 0.0094 | 0.936 |
| IE(0.33, 0.67) | 0.0076 | 0.0099 | -0.3026 | 0.0068 | 0.0066 | 0.933 | 0.0076 | 0.0071 | 0.0658 | 0.0073 | 0.0069 | 0.946 |
| TE(0.33, 0.67) | 0.0131 | 0.0160 | -0.2137 | 0.0112 | 0.0110 | 0.947 | 0.0131 | 0.0113 | 0.1374 | 0.0097 | 0.0095 | 0.937 |
| OE(0.33, 0.67) | 0.0069 | 0.0086 | -0.2464 | 0.0055 | 0.0054 | 0.952 | 0.0069 | 0.0062 | 0.1014 | 0.0056 | 0.0053 | 0.924 |

[a] IPW = Mean of inverse probability weighted estimates; ESE = empirical standard error; ASE = average estimated standard error; CP = empirical coverage probability).

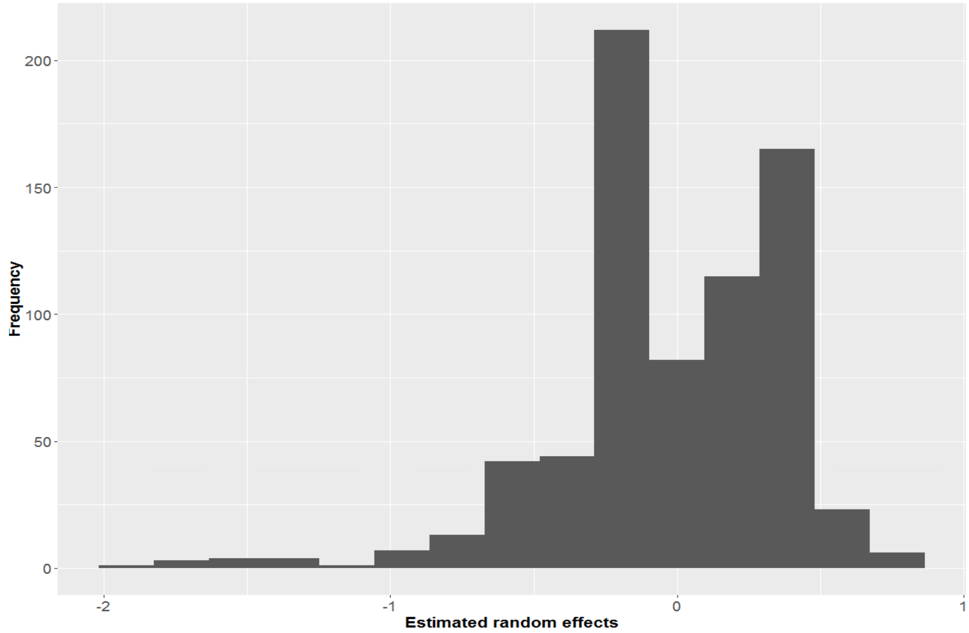


Figure 1: Distribution of the estimated random effects from the cluster-level propensity score model used in the analysis of disseminated effects in Optum's de-identified Clinformatics® Data Mart Database, 2010-2017, United States.

Table A4: International Classification of Diseases (ICD)-9/10-CM codes for opioid use disorder, opioid overdose and other comorbidities used in the analysis of disseminated effects in Optum's de-identified Clinformatics® Data Mart Database, 2010-2015, United States

| Condition | ICD Version | Code | Description |
|---|---|---|---|
| Opioid use disorder[1] | 9 | 304.00-304.03 | Opioid type dependence (unspecified; continuous; episodic) |
| | | 304.70-304.73 | Combinations of opioid type drug with any other drug dependence (unspecified; continuous; episodic) |
| | | 305.50-305.53 | Opioid abuse (unspecified; continuous; episodic) |
| Opioid overdose[2,3] | 9 | 965.0x | Poisoning by opiates and related narcotics |
| | | E850.0 - E850.2 | Accidental poisoning by heroin, methadone, or other opiates |
| | 10 | T40.0 | Poisoning by, adverse effect of and underdosing of opium |
| | | T40.1 | Poisoning by and adverse effect of heroin |
| | | T40.2 | Poisoning by, adverse effect of and underdosing of other opioids |
| | | T40.3 | Poisoning by, adverse effect of and underdosing of methadone |
| | | T40.6[4] | Poisoning by, adverse effect of and underdosing of other and unspecified narcotics |
| Depression[5] | 9 | 296.2 | Major depressive disorder single episode |
| | | 296.3 | Major depressive disorder recurrent episode |
| | | 296.5 | Bipolar i disorder, most recent episode (or current) depressed |
| | | 300.4 | Dysthymic disorder |
| | | 309 | Adjustment reaction |
| | | 311 | Depressive disorder, not elsewhere classified |

[1] Olfson, M., Wall, M., Wang, S., Crystal, S., and Blanco, C. (2018). Service use preceding opioid-related fatality. *American Journal of Psychiatry*, 175(6), 538-544..

[2] Frazier, W., Cochran, G., Lo-Ciganic, W. H., Gellad, W. F., Gordon, A. J., Chang, C. C. H., and Donohue, J. M. (2017). Medication-assisted treatment and opioid use before and after overdose in Pennsylvania Medicaid. *JAMA*, 318(8), 750-752.

[3] Daly, E. R., Dufault, K., Swenson, D. J., Lakevicius, P., Metcalf, E., and Chan, B. P. (2017). Use of emergency department data to monitor and respond to an increase in opioid overdoses in New Hampshire, 2011-2015. *Public Health Reports*, 132(suppl), 73S-79S.

[4] Excluding T406.06 and T40.696.

[5] Tonelli, M., Wiebe, N., Fortin, M., Guthrie, B., Hemmelgarn, B. R., James, M. T., ... and Sargious, P. (2016). Methods for identifying 30 chronic conditions: application to administrative data. *BMC Medical Informatics and Decision Making*, 15(1), 1-11.
